# Optimizing Screening for Intrauterine Fetal Growth Restriction in Low-Resource Settings Using 2D Ultrasound: A Deep Learning Approach

**DOI:** 10.64898/2026.05.04.26352354

**Authors:** Alfred Enywaku, Roy Alia Asiku

## Abstract

Severe fetal growth restriction (sFGR) affects 5 to 10% of pregnancies worldwide and is a major contributor to perinatal morbidity and mortality, particularly in low- and middle-income countries (LMICs). Traditional 2D ultrasound detection methods suffer from operator dependency, gestational age uncertainty, and limited access to Doppler in many low-resource facilities. This study presents a deep learning framework for sFGR screening and triage using 2D fetal abdominal ultrasound images designed to operate independently of precise gestational dating. Growth restriction severity labels were derived by mapping abdominal circumference measurements to INTERGROWTH-21st term percentiles as a gestational-age-normalized proxy for fetal size restriction when case-level gestational age or birth-weight data are unavailable. A systematic literature review of 37 studies revealed gaps in severity stratification and generalizability. We implemented a DenseNet-121-based model with abdominal circumference measurement for severity-aware classification using a retrospective single-center dataset of 1588 annotated fetal abdominal images from 169 term pregnancies. Patient-wise 3-fold cross-validation and ensemble testing yielded 93.7% accuracy, a weighted F1-score of 0.76, and ROC AUC ≥ 0.98 per class on heldout data. The approach outperforms previously reported single-center methods on this dataset while explicitly targeting LMIC-specific constraints. It demonstrates potential as a gestational-age-independent first-line triage layer for equitable prenatal screening, subject to prospective multi-site validation.

## 1 Introduction

Severe fetal growth restriction (sFGR), also known as intrauterine growth restriction (IUGR), represents a critical obstetric complication in which the fetus fails to achieve its genetic growth potential [Kamphof et al., 2022]. This condition affects approximately 3 to 10% of pregnancies globally, with a disproportionately higher burden in low- and middle-income countries (LMICs), where it contributes to up to 20% of newborns being small for gestational age [World Health Organization, 2019]. The consequences of sFGR are profound, and include increased risks of preterm birth, admission to the neonatal intensive care unit (NICU), and long-term complications such as neurodevelopmental impairment, cardiovascular disease, and metabolic syndrome [Lubrano et al., 2022, Malhotra et al., 2019, D’Agostin et al., 2023].

The conventional detection methods rely on 2D ultrasound biometry, where parameters such as abdominal circumference (AC) and estimated fetal weight (EFW) are measured and compared with gestational age (GA)-specific growth charts [Chew and Verma, 2023, Tongsong et al., 1999]. In LMICs, these approaches face several structural limitations, including high operator variability, inconsistent image-plane selection, and unreliable GA estimation due to late antenatal presentation or uncertain menstrual history [Dudley, 2021, Dahab and Sakellariou, 2020]. In sub-Saharan Africa, IUGR contributes to approximately 200 000 perinatal deaths annually. This burden is exacerbated by limited access to skilled sonographers, Doppler-capable systems, and advanced imaging platforms [Applegate et al., 2021].

Deep learning has emerged as a powerful tool in medical imaging. It provides automated feature extraction and classification capabilities that can mitigate some of these challenges [Alzubaidi et al., 2021, Mall et al., 2023]. Recent work in fetal ultrasound has reported promising performance in automated biometric measurement, standard plane detection, and anomaly screening [Chen et al., 2021, Yousefpour Shahrivar et al., 2023]. However, the majority of existing models still depend on accurate GA information, and few are designed to explicitly stratify disease severity, which limits their clinical usefulness in resource-constrained environments [Pokaprakarn et al., 2022, Lee et al., 2023].

This study addresses these gaps by proposing a comprehensive deep learning framework for sFGR detection and severity stratification based solely on 2D fetal abdominal ultrasound images. The framework is independent of precise GA dating and serves as a screening and triage tool that prioritizes sensitivity and negative predictive value. It relies on relative biometric relationships and severity-aware classification to decide which pregnancies should be escalated for full Doppler and multi-parameter assessment. The work follows a design science methodology (DSM) [Hevner et al., 2004], in which a technical artifact is constructed, evaluated, and refined in response to a clearly specified clinical problem.

The main contributions of this work are:

- A systematic PRISMA-guided review that maps the current landscape of deep learning approaches for sFGR detection and highlights unresolved limitations.
- A dual-measurement strategy for robust AC estimation that combines contour-based perimeter tracing with confidence-weighted ellipse fitting.
- A gestational-age-normalized severity labeling strategy that uses abdominal circumference-based percentiles at term as a pragmatic proxy for fetal size restriction when reliable GA or birth-weight data are unavailable.
- A classifier trained with patient-wise validation and ensemble testing, yielding strong performance on a publicly available clinical dataset.
- A severity-aware evaluation that emphasizes negative predictive value and clinically relevant operating points for use in LMIC screening workflows.

The remainder of this paper is organized as follows. Section 2 reviews related work and identifies key open challenges. Section 3 describes the dataset, preprocessing steps, feature extraction, labeling strategy, and model development. Section 4 presents the experimental results. Section 5 discusses clinical and methodological implications, as well as limitations and future extensions. Section 6 concludes.

## 2 Related Work

A systematic literature review was conducted following PRISMA guidelines to identify state-of-the-art deep learning models for sFGR diagnosis using 2D ultrasound images. Searches were performed in IEEE Xplore, ScienceDirect, PubMed, and Google Scholar between 2009 and 2024 using MeSH terms and keywords such as “fetal growth restriction,” “intrauterine growth restriction,” “2D ultrasound,” and “deep learning”, including spelling variants and related phrases.

From 548 initial records, 37 studies met the inclusion criteria after duplicate removal, title and abstract screening, and full-text assessment. Eligible studies were peer-reviewed, made use of 2D ultrasound, and reported some form of deep learning-based analysis for FGR-related diagnosis or surrogate tasks. Reviews and studies based on non-2D modalities, such as MRI or 3D ultrasound, and work unrelated to FGR were excluded.

Overall, 48.7% of the selected studies used CNN-based architectures, 35.1% used ensemble models, and 16.2% used hybrid approaches. Approximately 78% trained on clinically annotated datasets acquired during routine prenatal care, while 16.2% used the HC18 challenge dataset [van den Heuvel et al., 2018].

From the analysis, a few key observations became clear.

### Biometric Measurement

A large fraction of studies focused on automated measurement of head, abdominal, or femoral biometrics for GA estimation or growth assessment [Zeng et al., 2021, Sobhaninia et al., 2019]. Płotka et al. [2021] proposed a multi-task architecture that performs segmentation and biometric measurement. They achieved 95.6% classification accuracy and Dice scores between 0.90 and 0.92 for fetal structures.

### Plane Classification

Other work addressed the automatic recognition of standard planes. Burgos-Artizzu et al. [2020] used ResNet50 to classify maternal-fetal planes and reported 95.3% accuracy. They demonstrated that deep models can approximate expert-level view selection.

### Quality Assessment

Deep learning has also been applied to the assessment of image quality and adherence to clinical standards. Wu et al. [2017] reported 85.7% agreement with expert readers when grading image quality in obstetric ultrasound. This work supports the use of AI as a quality gate before downstream measurements.

### FGR-Specific Detection

A smaller subset of studies explicitly targeted FGR. Dong et al. [2025] trained a model on placental images to detect early FGR and achieved an F1-score of 0.765 with an AUC of 0.80. Mikołaj et al. [2025] reported improved sensitivity for small-for-gestational-age (SGA) fetuses, with SGA detection sensitivity reaching 70% compared to 58% achieved by the conventional Hadlock formula, using a deep learning model trained on over 433,000 ultrasound images.

### Non-Image Modalities

For context, related work using non-image signals has also achieved high performance. Pini et al. [2021] used fetal heart rate time series and obtained 93% classification accuracy, while Rescinito et al. [2023] synthesized evidence across modalities and reported pooled sensitivity of 0.84 and specificity of 0.87 for machine learning-based approaches.

Across these studies, several common limitations were identified. First, many models are tightly coupled to gestational age and rely on precise GA estimates during training and inference [Pokaprakarn et al., 2022]. Second, few works explicitly stratify severity levels of growth restriction, instead collapsing outcomes into binary normal versus abnormal classes. Third, there is limited evidence of external validation or multi-center generalizability [Sendra-Balcells et al., 2023]. The framework presented in this paper is designed to address these issues by providing GA-independent, severity-aware classification on a publicly available dataset while explicitly acknowledging the single-center nature of the current study.

## 3 Materials and Methods

The study followed the DSM paradigm [Hevner et al., 2004], with phases covering problem identification, literature review, model design, and empirical evaluation. In this section, we describe the dataset, preprocessing pipeline, feature extraction and labeling, model development, and evaluation protocol.

### 3.1 Dataset Acquisition and Characteristics

The Fetal Abdominal Structures Segmentation Dataset [Da Correggio et al., 2024] was selected after a comprehensive search of open repositories such as Zenodo, Mendeley Data, and Kaggle. The dataset contains 1588 ultrasound images from 169 term pregnancies, with a mean of 9.4 images per patient (Fig. 1). Images were acquired between 2021 and 2023 at the University Hospital Polydoro Ernani de São Thiago in Brazil using Siemens Acuson, Voluson 730, and Philips EPIQ Elite scanners with 2 to 9 MHz transducers. Expert annotators provided structure-wise masks for the aorta, vein, stomach, and liver. The cohort includes both normal and complicated pregnancies, including IUGR, diabetes, and preeclampsia. Ethical approval for data collection was obtained from the UFSC ethics committee. A representative abdominal image is shown in Fig. 2.

**Figure 1.**
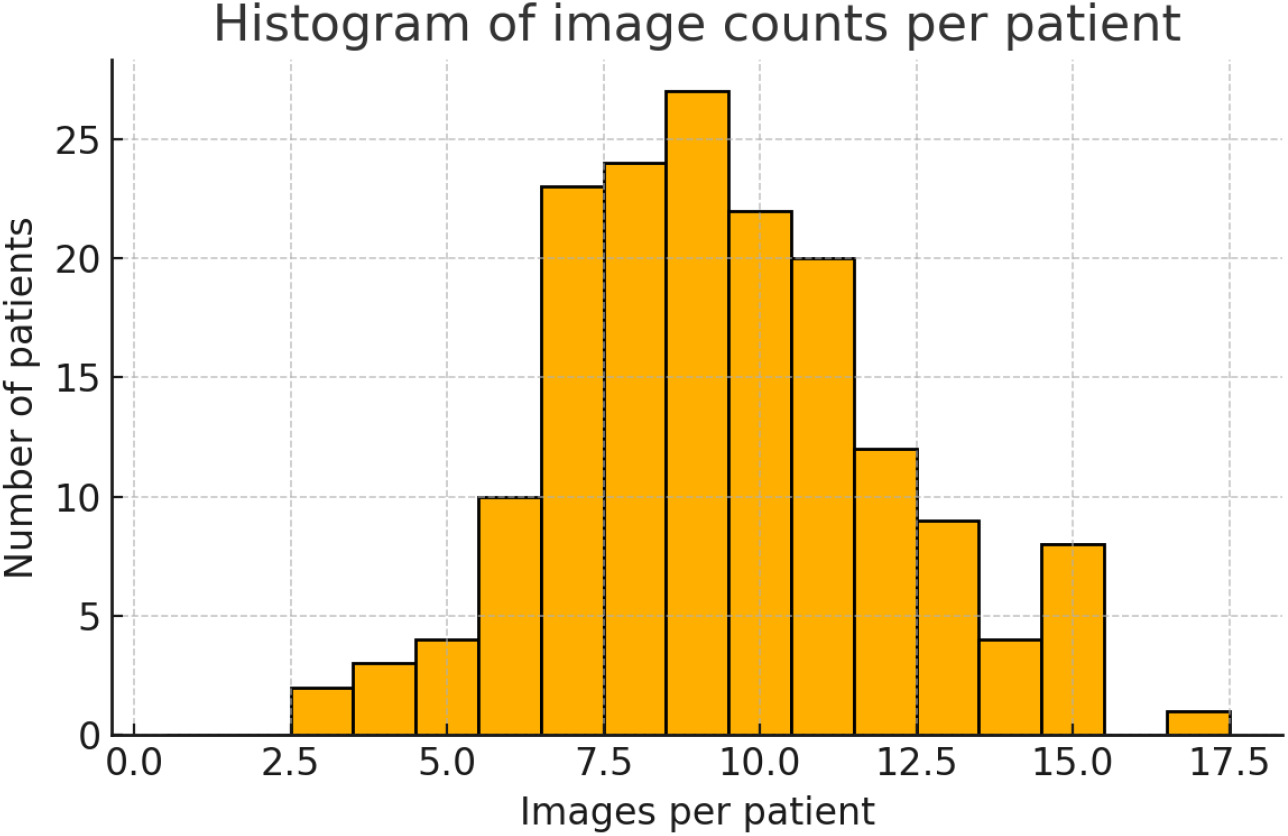
Distribution of image counts per patient in the fetal abdominal dataset.

**Figure 2.**
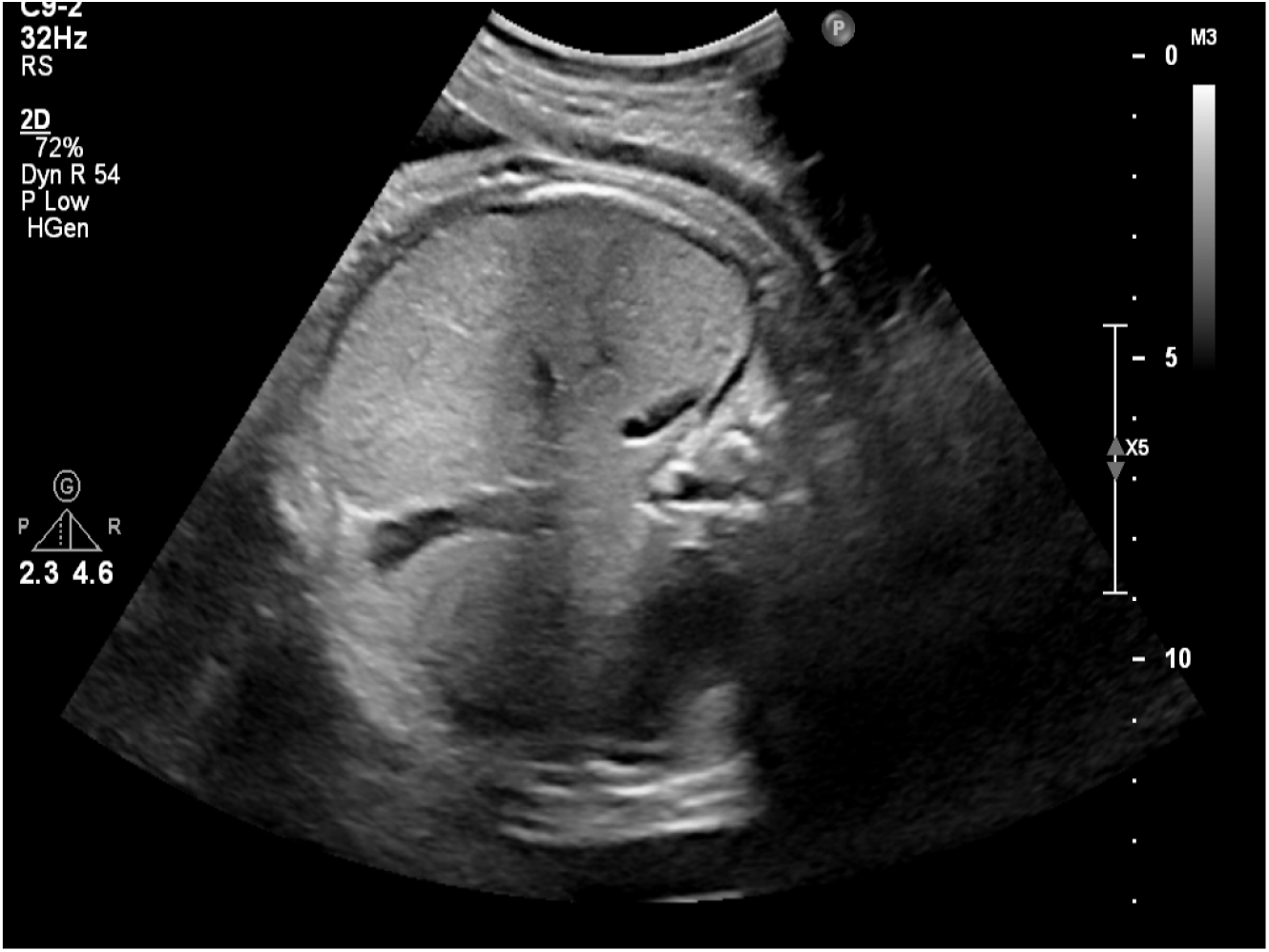
Example fetal abdominal ultrasound image from the dataset.

Because all images originate from a single tertiary referral center and a limited set of scanner models, the present study should be interpreted as a proof-of-concept evaluation of the proposed framework on this dataset. External validation on independent cohorts from other regions and care levels is required before clinical deployment.

### 3.2 Data Preprocessing

Data preprocessing is a critical step in preparing ultrasound images for deep learning analysis, since raw medical images often contain artifacts, varying formats, and acquisition inconsistencies that can adversely affect model performance [Alhubaidi et al., 2021]. The preprocessing pipeline was designed to standardize inputs while preserving clinically relevant information.

From the input images, the pixel array was extracted with PyDICOM, converted to 8-bit grayscale, and then to BGR format. This normalization ensured a consistent intensity range across the dataset. Text overlays and scale bars are common artifacts in clinical ultrasound. They were removed to prevent the network from learning spurious correlations. Text detection was done using adaptive thresholding followed by morphological operations with horizontal (25 × 1) and vertical (1 × 5) kernels. Scale bar detection focused on the lateral 12% of the image width where bars are typically located. It was done using vertical morphology to isolate elongated structures. The detected regions were then inpainted using OpenCV’s TELEA algorithm with a 5-pixel radius, which fills the masked areas with contextually plausible values derived from surrounding pixels.

To preserve the anatomical context, the entire scan sector was retained without cropping or zero-filling. This design choice avoided loss of peripheral information that might contribute to robust feature learning and contrasts with aggressive cropping strategies that may discard useful structures [Sendra-Balcells et al., 2023].

To increase robustness to acquisition variability and to mitigate the relatively limited dataset size, we applied data augmentation with the Albumentations library. Applied geometric transformations included random rotations at 90 degrees with probability *p* = 0.5 to promote orientation invariance, horizontal and vertical flips with *p* = 0.5 to exploit anatomical symmetry, and Shift Scale Rotate with shift_limit=0.1, scale_limit=0.2, rotate_limit=20°, and *p* = 0.7 to simulate differences in probe position and fetal posture. Elastic Transform with parameters *α* = 120, *σ* = 6, and *p* = 0.3 was used to approximate tissue deformation patterns common in ultrasound imaging.

Pixel-level augmentations were also applied to address intensity and noise variability. Random Brightness Contrast with limit=0.2 and *p* = 0.5, together with Random Gamma with range 80 to 120 and *p* = 0.3, simulated different gain and dynamic range settings. Gaussian Noise with variance 10 to 50 and *p* = 0.3 approximated acoustic noise, and Contrast Limited Adaptive Histogram Equalization (CLAHE) with clip_limit=2.0 and *p* = 0.4 enhanced local contrast to improve visibility of soft-tissue boundaries in low-quality scans. Together, these operations were chosen to refiect realistic perturbations in LMIC settings where scanner quality and maintenance vary [Dahab and Sakellariou, 2020]. A preprocessed image sample is shown in Fig. 3.

**Figure 3.**
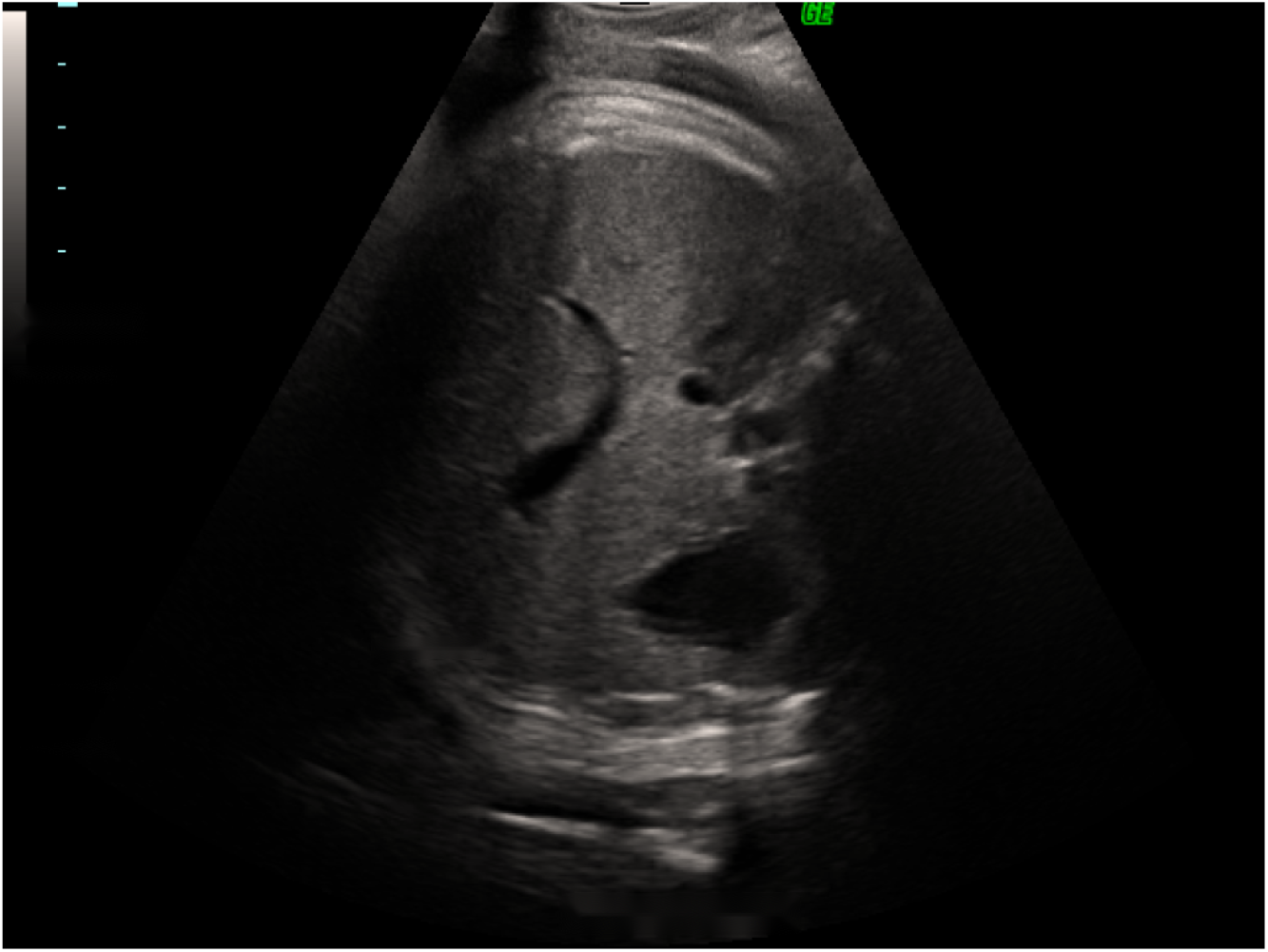
Illustration of preprocessing steps applied to a raw abdominal ultrasound image.

### 3.3 Feature Extraction

Feature extraction focused on robust AC estimation, which is a key biometric marker for IUGR assessment [Tongsong et al., 1999]. To reduce dependence on a single geometric assumption, we implemented a dual-method strategy. The first method used contour-based perimeter tracing to capture detailed abdominal outlines, while the second relied on ellipse fitting to provide a standardized approximation that is more tolerant of partial occlusions and local mask imperfections. Both methods operated on unified masks derived from expert annotations of the artery, vein, stomach, and liver.

#### 3.3.1 Perimeter-Based Method

The perimeter-based method aimed to obtain a precise boundary of the abdominal region, which is particularly valuable in cases where the shape deviates from an ideal ellipse. Individual binary masks for the artery (*M*_*art*_), liver (*M*_*liv*_), stomach (*M*_*sto*_), and vein (*M*_*vei*_) were combined into a single abdominal mask by logical OR operation:

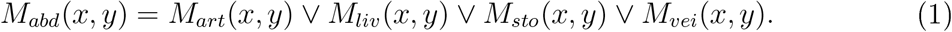

We then applied a sequence of refinement steps. First, internal voids were filled using `ndimage.binary_fill_holes`, yielding *M*_*filled*_. Small isolated components were removed with `skimage.remove_small_objects`. This suppressed noise while preserving the main abdominal contour. Gaussian smoothing with a 5 × 5 kernel, followed by re-binarization, reduced jagged edges. Finally, morphological closing with a 5 × 5 elliptical structuring element connected nearby regions and closed narrow gaps:

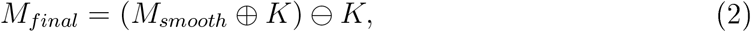

where *K* denotes the structuring element, and ⊕ and ⊖ indicate dilation and erosion, respectively.

Contours were extracted from *M*_*final*_ using `cv2.findContours` with the `CHAIN_APPROX_SIMPLE` contour approximation method. This produced a point set 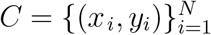. The contour perimeter in pixels was computed as

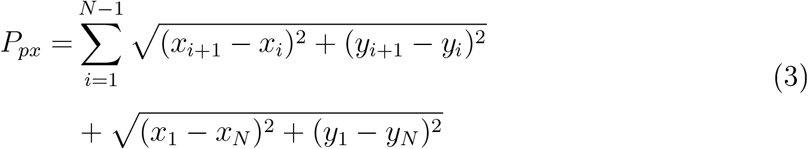

and converted to millimeters using the pixel-to-millimeter ratio *R*_*px/mm*_ = 0.28:

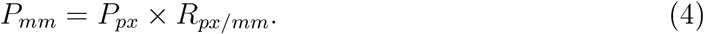

#### 3.3.2 Ellipse Fitting Methods

Ellipse-based methods provide a compact representation of the abdominal contour and are often more robust to local segmentation artifacts. We implemented three variants.

The direct fit variant applied `cv2.fitEllipse` to the largest contour, obtaining semi-axes *a* and *b*. The circumference was approximated using Ramanujan’s formula [Yillarino, 2005]:

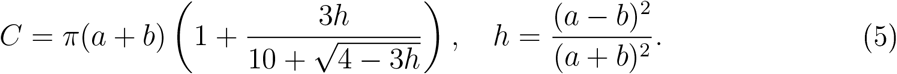

The active contour variant first refined the contour with the `skimage.active_contour` algorithm [Kass et al., 1988], which evolves an initial snake towards prominent edges, and then fit an ellipse to the refined curve using the same approximation for circumference.

The moment-based variant constructed a covariance matrix from the central moments of the mask and used its eigenvalues to derive the ellipse axes. Let *λ*_1_ and *λ*_2_ be the eigenvalues of the covariance matrix; then

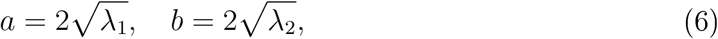

and the circumference was again estimated with Ramanujan’s formula.

Each ellipse estimate was assigned a confidence score. For the direct and active-contour methods, the score combined the intersection-over-union (IoU) between the ellipse and the mask with an aspect-ratio term. For the moment-based approach, the score combined low eccentricity with a high area ratio between mask and ellipse. The method with the highest confidence was selected as the final ellipse-based AC measurement. An illustration of the fitted ellipses overlaid on the abdominal mask is shown in Fig. 4.

**Figure 4.**
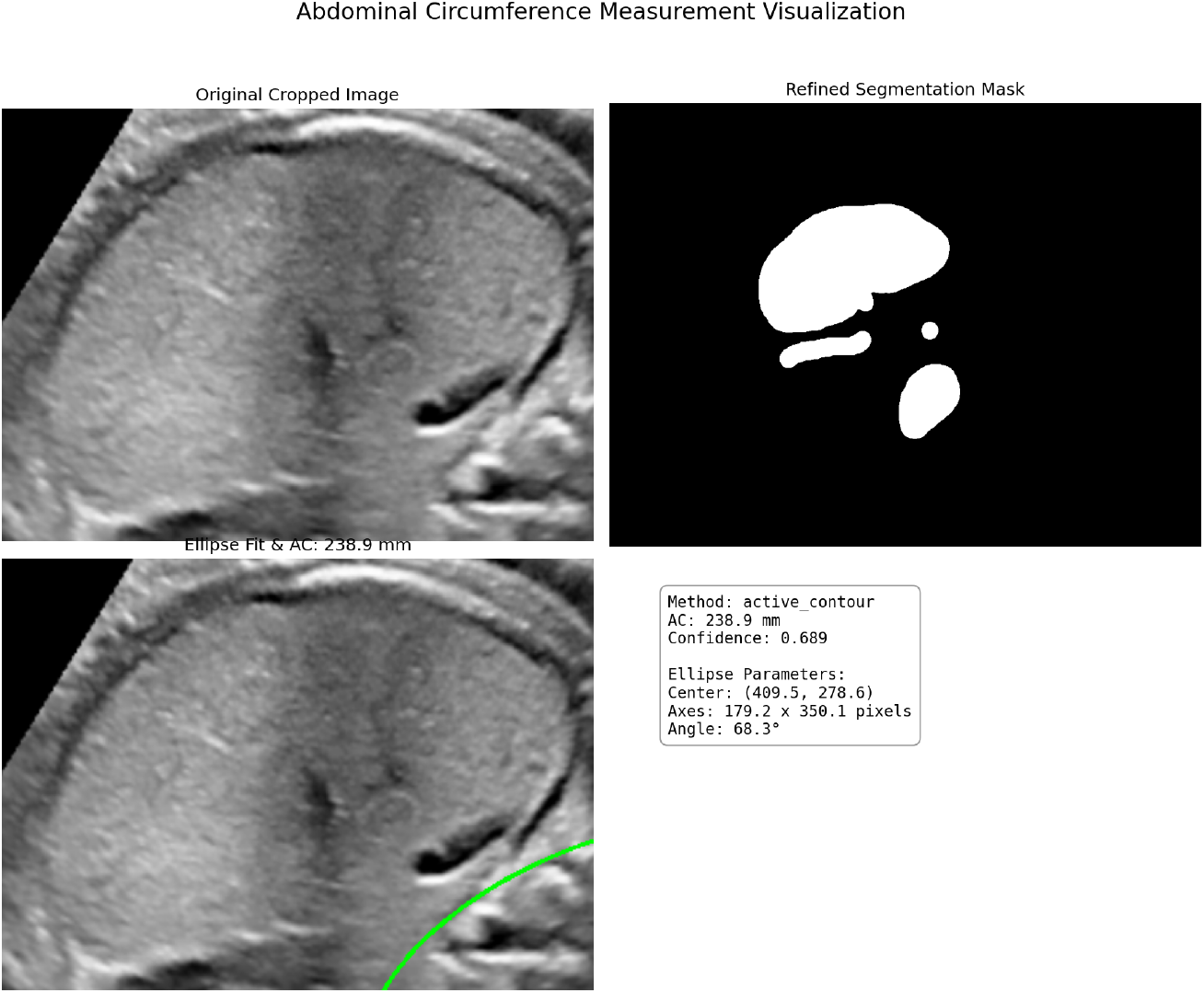
Visualization of abdominal circumference measurement from unified masks using perimeter tracing and ellipse fitting.

Unless otherwise specified, “ellipse-based AC” in the remainder of the paper refers to this confidence-weighted circumference estimate. This ellipse-based AC was used for percentile mapping and severity labeling, whereas the perimeter-derived AC was retained for descriptive analysis of measurement behaviour and to characterize the distributional differences between methods (Section 4.1). As a result, extreme upper-tail perimeter values do not directly influence the class labels supplied to the classifier.

### 3.4 Labeling

To assign clinically meaningful labels under the constraint that reliable per-case GA, EFW, and neonatal outcomes were not available, we mapped abdominal circumference measurements to severity categories at term using established growth standards. In particular, we focused on images acquired around 37 weeks of gestation and referenced the INTERGROWTH-21st growth charts [Stirnemann et al., 2020]. Operational clinical definitions of FGR often rely on thresholds such as EFW or AC below the 10th, 5th, or 3rd percentile for GA at delivery [Chew and Verma, 2023, Tongsong et al., 1999, Stirnemann et al., 2020]. By mapping all cases to a common term reference frame, we preserve this percentile-based notion of severity while removing the need for precise GA at the time of scan, which is frequently unavailable in retrospective LMIC data.

For a reference gestational age of 37 weeks, we approximated the 10th, 5th, and 3rd percentiles as follows: 10th percentile AC of 290 mm and EFW of 2500 g, 5th percentile AC of 285 mm and EFW of 2400 g, and 3rd percentile AC of 280 mm and EFW of 2300 g. Unless otherwise stated, “AC’ in this section refers to the confidence-weighted ellipse-based estimate obtained as described in Section 3.

Direct EFW measurements were not available in the dataset, so we implemented a proxy estimation based on AC. For AC values below the 3rd percentile, EFW was set to 95% of the 3rd percentile threshold. For intermediate AC values, EFW was obtained by linear interpolation between the corresponding percentile anchors. If AC was at or above the 10th percentile, EFW was approximated by the 50th percentile value of 3300 g.

We then applied a minimum-criteria rule to derive four severity classes: severe, moderate, mild, and normal. A case was labeled as severe if either AC or the estimated EFW fell below the 3rd percentile. If at least one of the two measurements was below the 5th percentile but not in the severe range, the case was labeled as moderate. Mild cases were defined by at least one measurement below the 10th percentile but not meeting the stricter thresholds for moderate or severe restriction. Finally, when both AC and estimated EFW were at or above the 10th percentile, the image was labeled as normal. This scheme emphasizes sensitivity to more severe restriction while preserving a clinically interpretable ordinal structure across classes.

This proxy-based labeling should be viewed as a principled approximation of guideline-defined severity under data limitations rather than as a substitute for true outcome-based labels. Future work will compare these proxy labels against outcome-based definitions in cohorts where reliable GA, EFW, and birth outcomes are available, in order to quantify any systematic bias introduced by the proxy.

### 3.5 Model Development

The classification model was built on a DenseNet-121 backbone [Hasan et al., 2021] (Fig. 5) chosen for its strong performance on medical imaging tasks and its dense connectivity pattern, which facilitates feature reuse and gradient flow. The convolutional trunk was initialized with ImageNet-pretrained weights and adapted to the fetal abdominal domain using a task-specific classification head.

**Figure 5.**
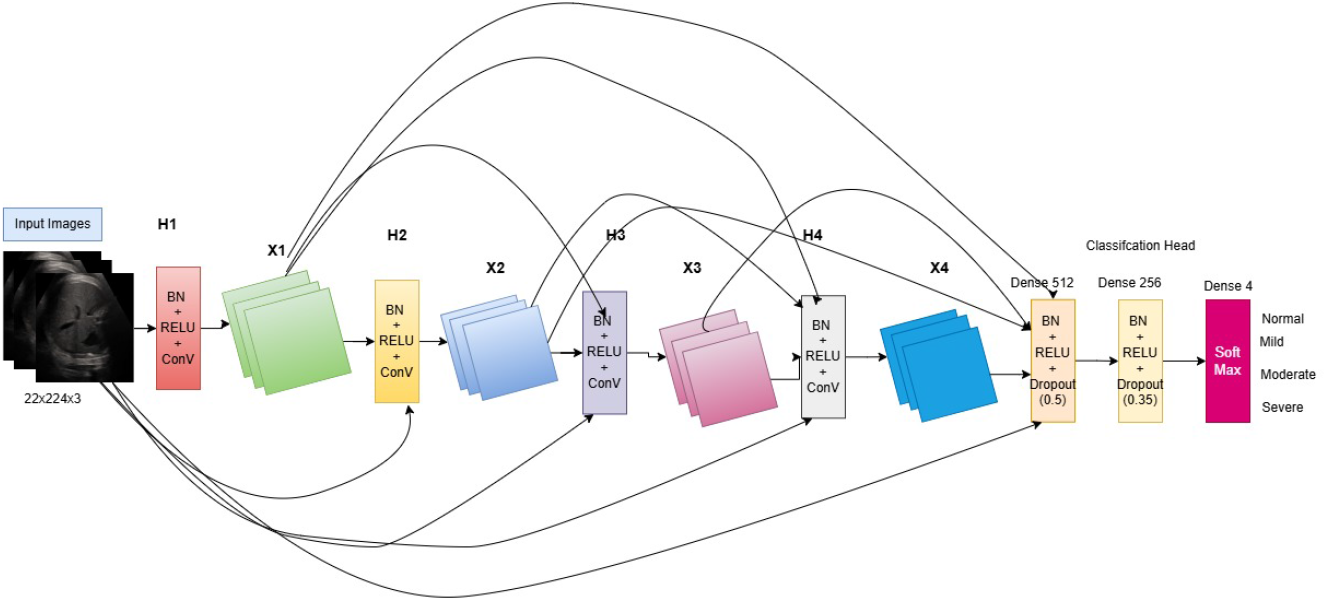
Adopted architecture based on DenseNet-121 pre-trained model.

The custom head consisted of a sequence of fully connected layers with batch normalization, non-linear activation, and dropout regularization. The DenseNet feature vector was first projected from 1024 to 512 units, followed by batch normalization, a ReLU activation, and a dropout rate of 0.5. A second projection from 512 to 256 units used the same pattern with a dropout rate of 0.35. The final linear layer mapped the 256-dimensional representation to four logits corresponding to the four severity classes.

To address class imbalance, we experimented with two loss functions. The primary objective was focal loss with class-specific weighting *α* and focusing parameter *γ* = 2.0, which down-weights easy examples and increases emphasis on misclassified or minority-class samples. As an alternative, we implemented a class-weighted cross-entropy loss where weights were inversely proportional to class frequencies. The results presented in this work were obtained using the class-weighted cross-entropy loss.

Training followed a two-phase strategy combining transfer learning and fine-tuning. In the first phase, all DenseNet layers were frozen and only the classification head was trained for 25 epochs. We used the AdamW optimizer with an initial learning rate of 10^−3^ and weight decay of 10^−4^. A cosine annealing learning rate scheduler with *T*_max_ = 25 and minimum learning rate of 10^−6^ was applied, and early stopping based on validation loss prevented overfitting.

In the second phase, the last three dense blocks of the backbone were unfrozen to allow feature adaptation to the ultrasound domain. The learning rate was reduced to 10^−5^, and we used a ReduceLROnPlateau scheduler monitoring the validation F1-score. Early stopping with the same patience settings ensured that training terminated once performance converged.

To obtain robust performance estimates, we used patient-wise 3-fold cross-validation. Patients, rather than images, were partitioned so that all images from a given patient were assigned to the same fold. Fifteen percent of patients were held out as an independent test set and not used during training or validation. The remaining patients were divided into three folds with approximately balanced class distributions. For final evaluation, the three models were combined in an ensemble: each model produced class probabilities on the test set, and the probabilities were averaged to obtain the final predictions. In the intended clinical use case, these probabilities can also be thresholded to prioritize high sensitivity and negative predictive value for screening.

### 3.6 Evaluation Metrics

Model performance was quantified using a set of standard classification metrics. Overall accuracy was defined as the proportion of correctly classified images across all four severity classes. For each class, we computed precision, recall, and the F1-score. Since screening tools are often used to rule out disease, we also report the negative predictive value (NPV), which indicates the probability that a predicted non-severe case is truly non-severe.

Discriminative ability was assessed using receiver operating characteristic (ROC) curves and the area under the ROC curve (AUC). For the multi-class setting, class-wise one-versus-rest curves were computed. Confusion matrices were constructed to visualize error patterns and misclassification between adjacent severity levels.

For the ensemble, we evaluated metrics both globally and per class on the held-out test set. Where appropriate, we compare these results qualitatively with those reported in recent literature on FGR detection with ultrasound.

## 4 Results

### 4.1 Feature Analysis

We first examined the abdominal circumference estimates produced by the perimeter-based and ellipse-based methods. The perimeter-derived AC values exhibited a broader dynamic range between 76 and 455 mm. This reflects the method s sensitivity to fine-grained contour variation and the accumulation of local irregularities (Fig. 6). In contrast, ellipse-based measurements were more conservative, spanning a narrower interval of 72 to 310 mm.

**Figure 6.**
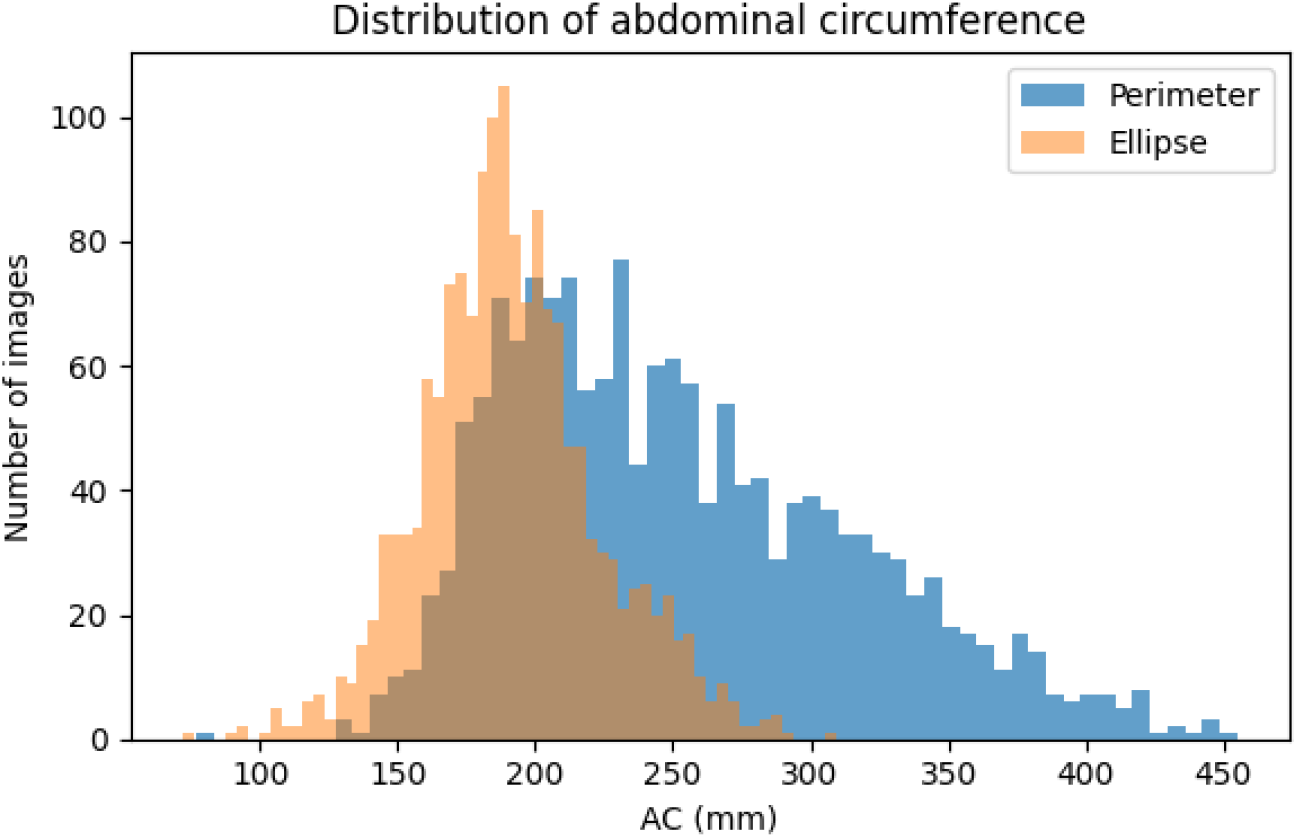
Distribution of abdominal circumference values obtained with perimeter tracing and ellipse fitting.

The small number of perimeter measurements above approximately 360 mm corresponded to masks with highly irregular outer borders. Because percentile-based severity labels are derived from the confidence-weighted ellipse measurement (Section 3), these perimeter outliers are not propagated directly into the class definitions. Instead, the perimeter distribution is used to characterize the behaviour of the segmentation pipeline and to highlight cases that may benefit from additional quality control in future prospective deployments.

A direct comparison between the two methods is shown in the methods confusion matrix (Fig. 7). In general, the ellipse method tended to slightly underestimate circumference in larger abdomens, which is consistent with approximating complex shapes by smooth ellipses. Box plots stratified by severity class (Fig. 8) indicated that both methods preserved the expected ordering of classes, with decreasing AC from normal to severe. The perimeter method accentuated separation between extremes, whereas the ellipse method produced tighter within-class distributions.

**Figure 7.**
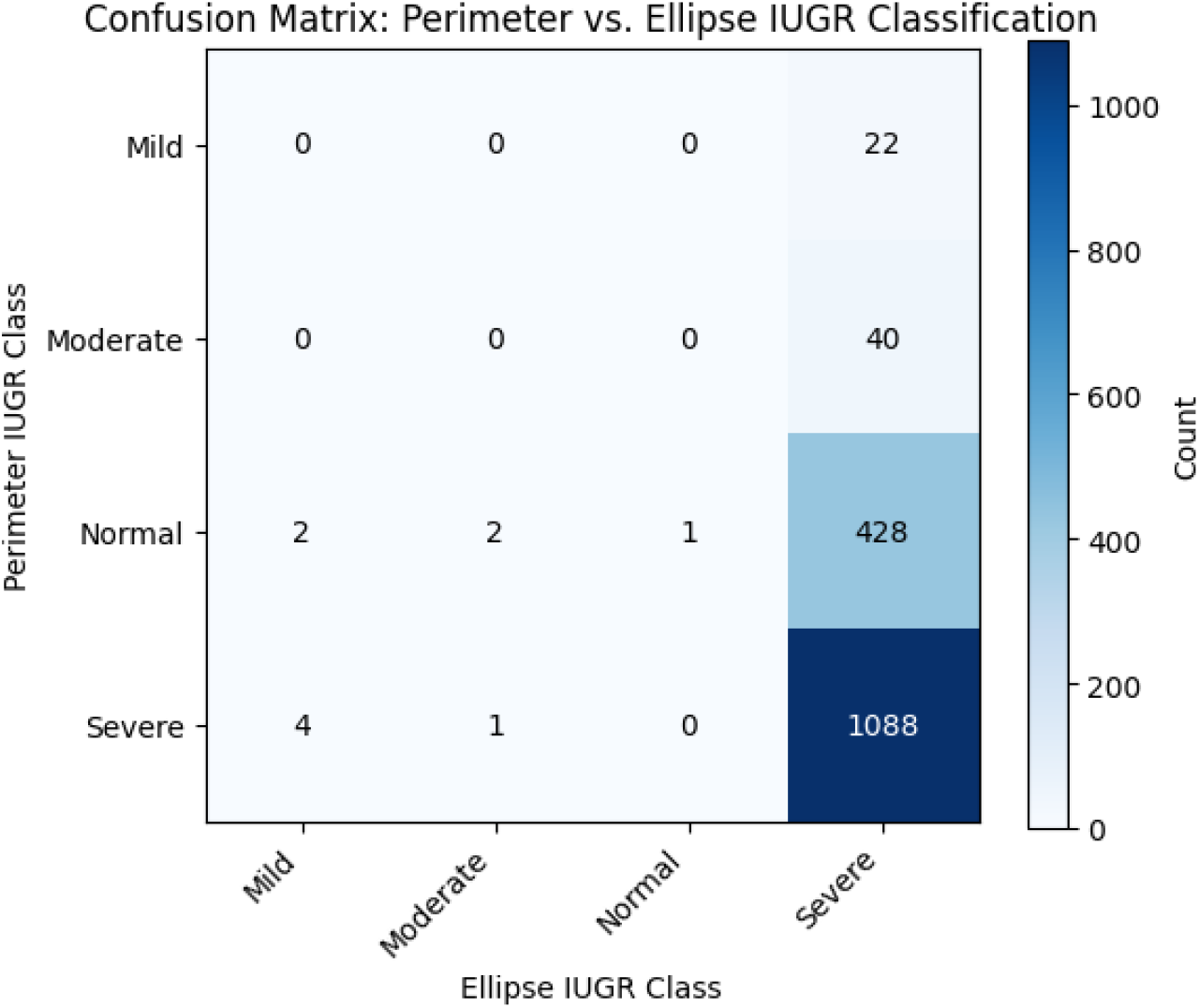
Confusion matrix comparing class assignments derived from perimeter-based and ellipse-based abdominal circumference estimates.

**Figure 8.**
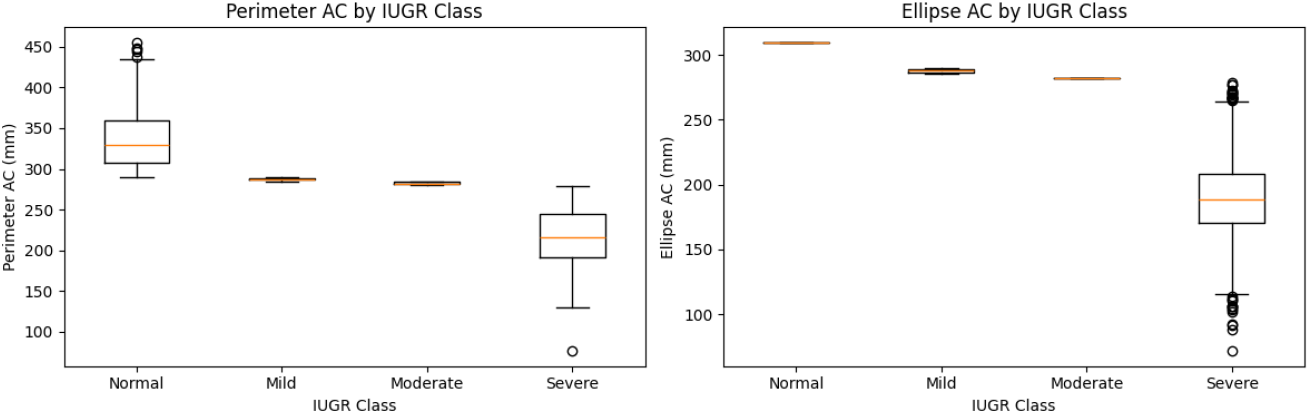
Abdominal circumference distributions per severity class for perimeter-based (left) and ellipse-based (right) measurements.

### 4.2 Training Results

Training curves for the three cross-validation folds showed rapid improvements in both training and validation metrics during the initial epochs, followed by a plateau phase. In each fold, validation loss decreased substantially in the first phase, confirming the benefit of transfer learning from the pretrained DenseNet backbone. The fine-tuning produced further, more moderate gains in F1-score and AUC.

The cross-validation confusion matrices (Figs. 9 to 11) revealed that most errors occurred between neighboring severity levels, which is expected given the ordinal nature of the labels. In the first fold, intermediate classes tended to collapse towards the normal and severe extremes, suggesting that the model initially favored clearer patterns at the tails of the distribution. In folds 2 and 3, better calibration of decision boundaries led to more balanced performance across the four classes.

**Figure 9.**
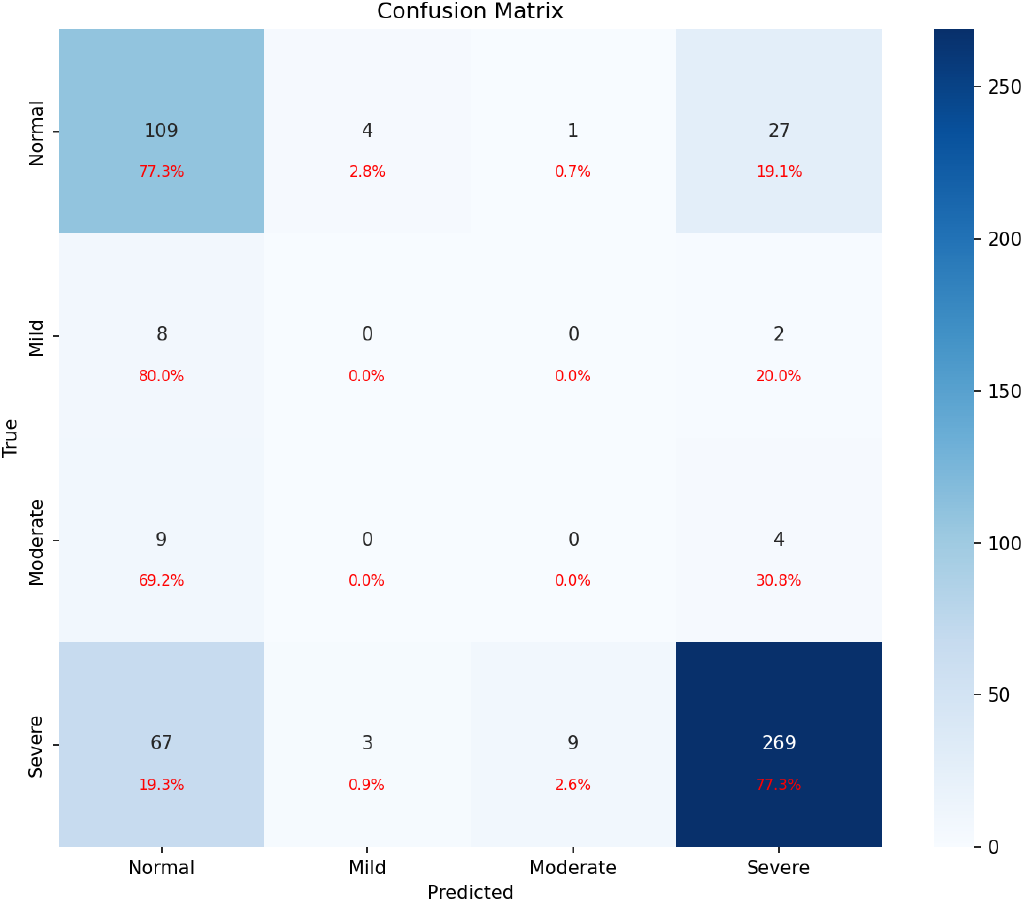
Cross-validation confusion matrix for fold 1.

**Figure 10.**
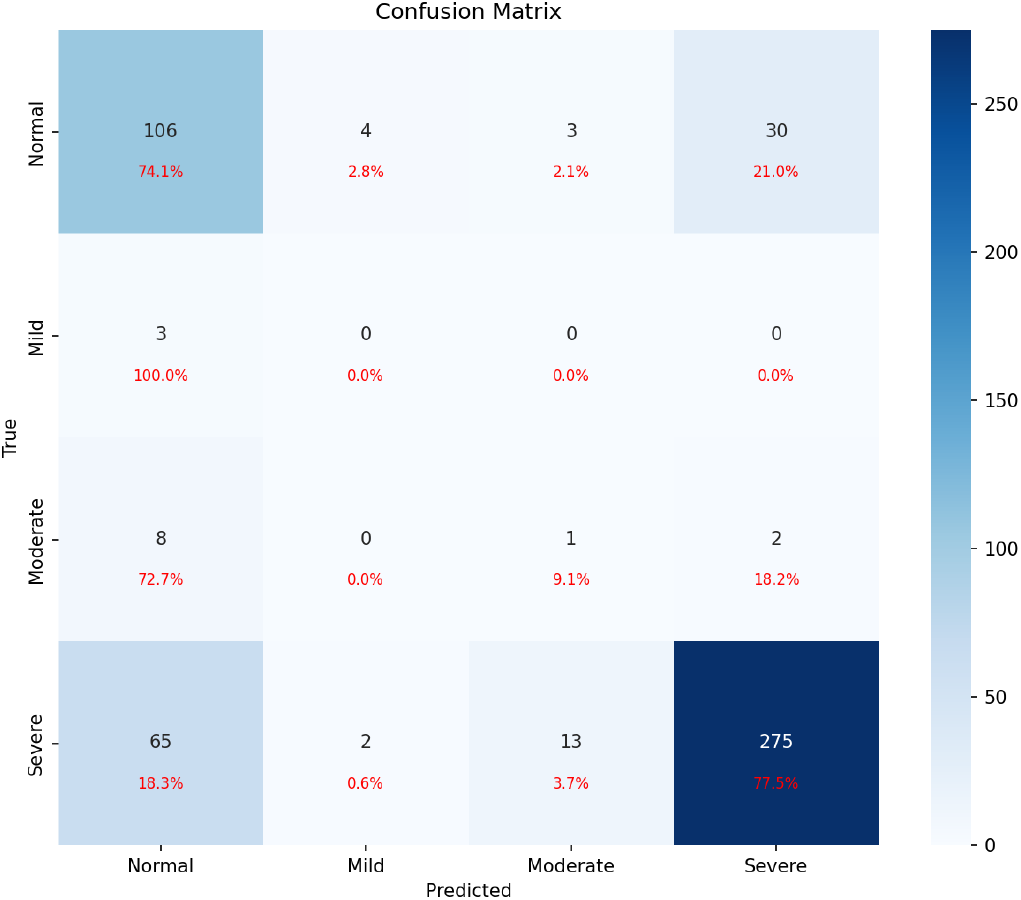
Cross-validation confusion matrix for fold 2.

**Figure 11.**
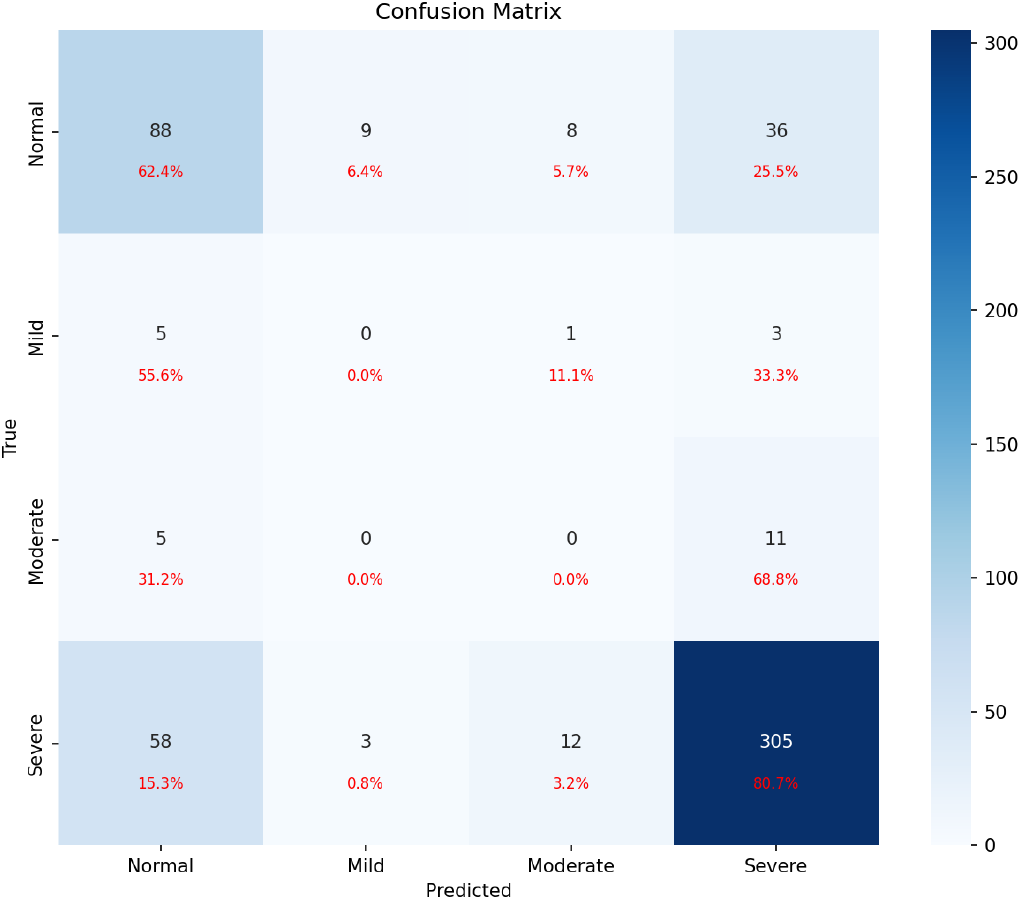
Cross-validation confusion matrix for fold 3.

The corresponding ROC curves (Figs. 12 to 14) demonstrated strong discrimination for the normal and severe classes, with AUC values above 0.80. The mild and moderate classes, which are naturally harder to distinguish due to overlapping AC ranges, exhibited lower AUC values in the range 0.46 to 0.60 during cross-validation. This pattern motivated the use of ensemble testing and severity-aware interpretation on the final test set.

**Figure 12.**
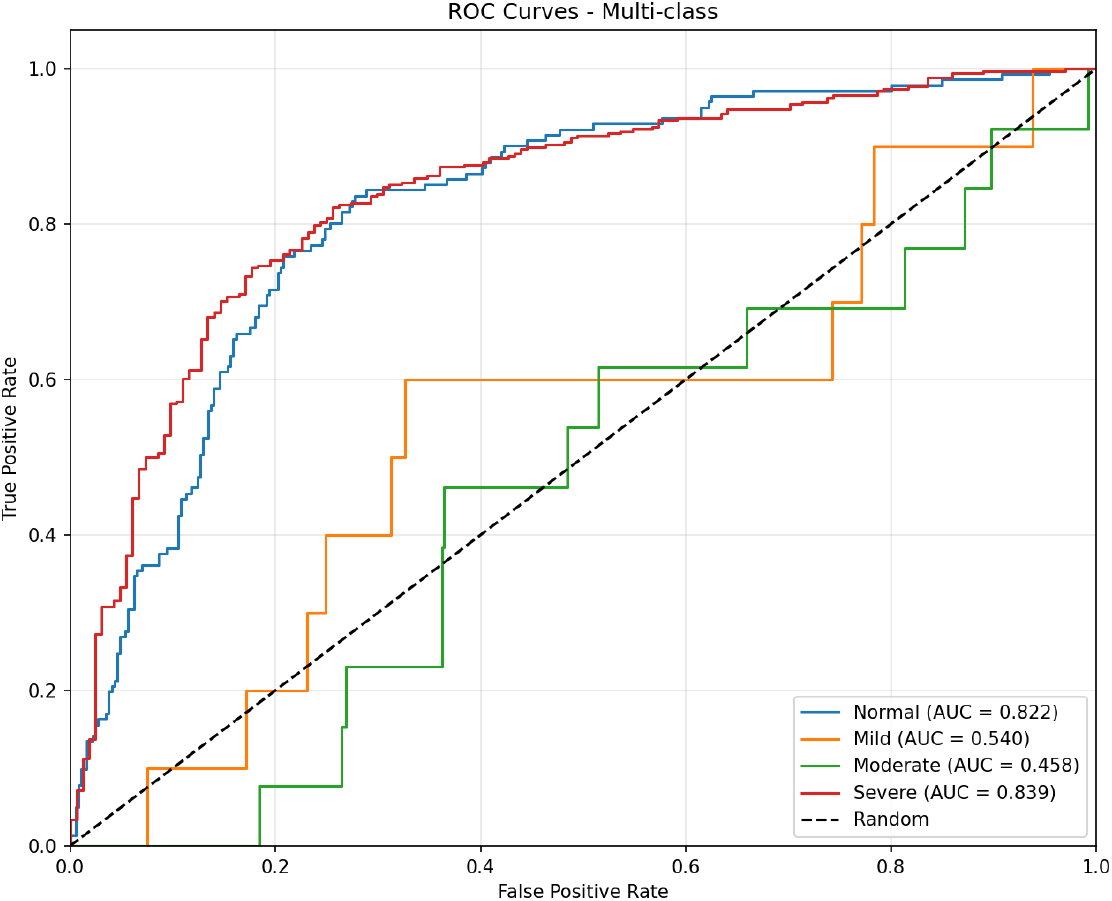
Per-class ROC curves for fold 1.

**Figure 13.**
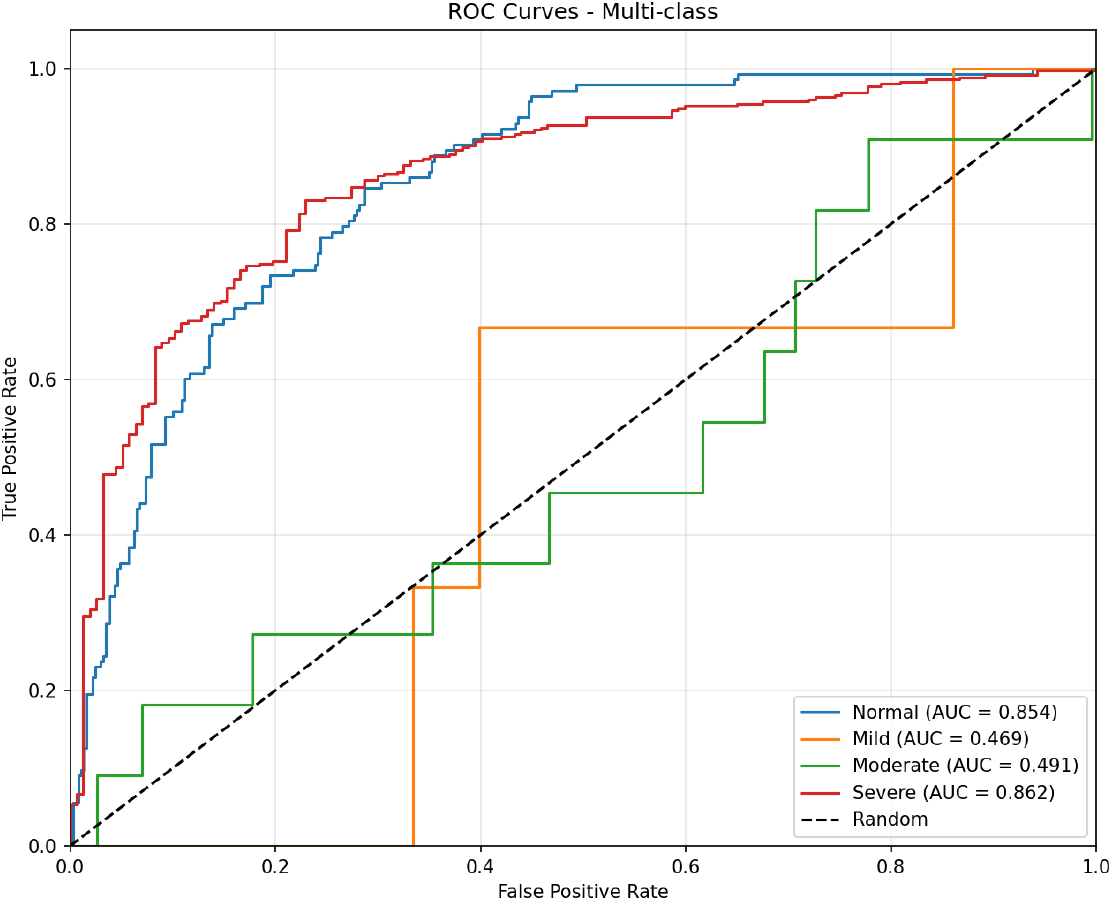
Per-class ROC curves for fold 2.

**Figure 14.**
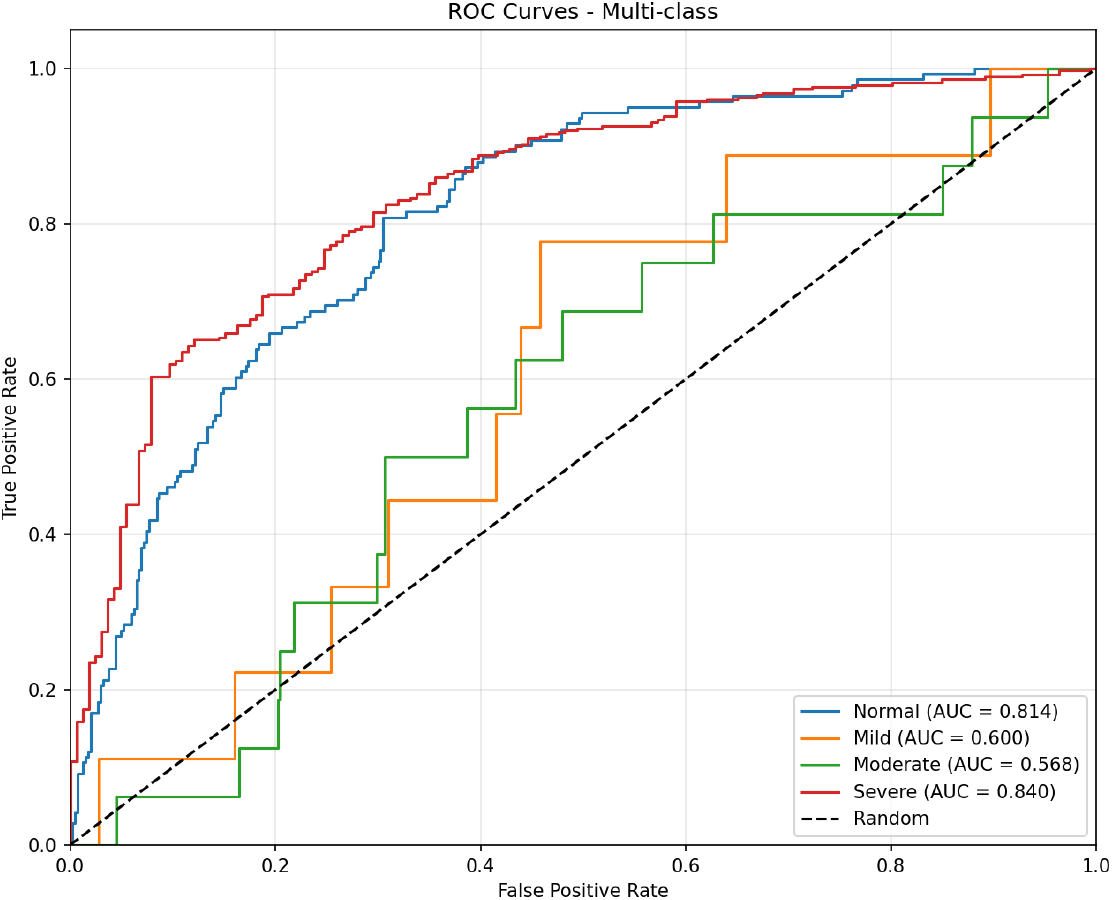
Per-class ROC curves for fold 3.

### 4.3 Test Set Evaluation

On the held-out test set, the ensemble model achieved an overall accuracy of 93.7% and a weighted F1-score of 0.76. The mild and moderate categories were predicted with particularly high F1-scores. This shows that the combination of dual AC measurement and severity-aware labeling produced stable intermediate classes. The normal and severe classes also achieved high precision and recall. This confirms that the system can reliably distinguish clinically most critical extremes.

The confusion matrix for the test set (Fig. 15) shows that misclassifications were largely confined to adjacent categories, with very few normal cases predicted as severe or vice versa. This behavior is desirable in a screening tool, since it reduces the risk of gross misclassification with major clinical consequences.

**Figure 15.**
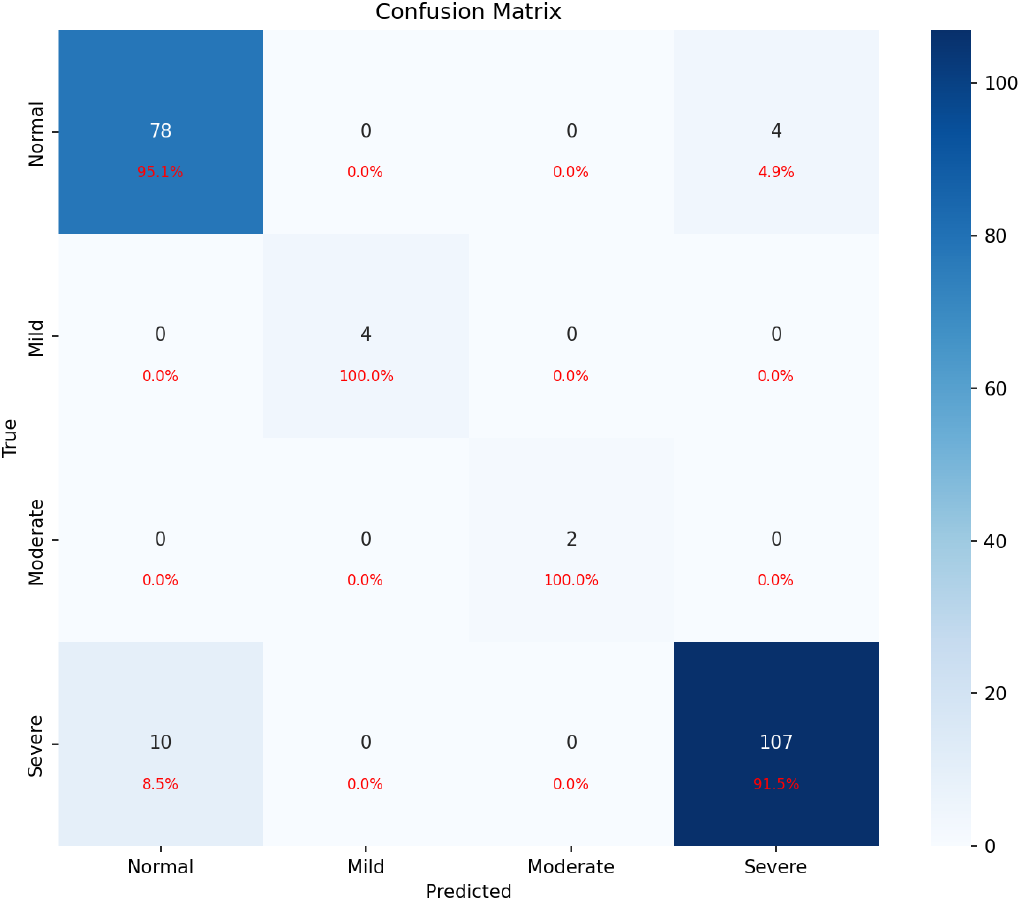
Confusion matrix for the ensemble model on the held-out test set.

Per-class ROC curves (Fig. 16) yielded AUC values of at least 0.98 for all four classes. It indicates excellent discrimination when using probability thresholds instead of hard class assignments. This supports the use of the model not only for point predictions but also as a probabilistic triage tool in clinical workflows where decision thresholds can be tuned to prioritize sensitivity and NPV.

**Figure 16.**
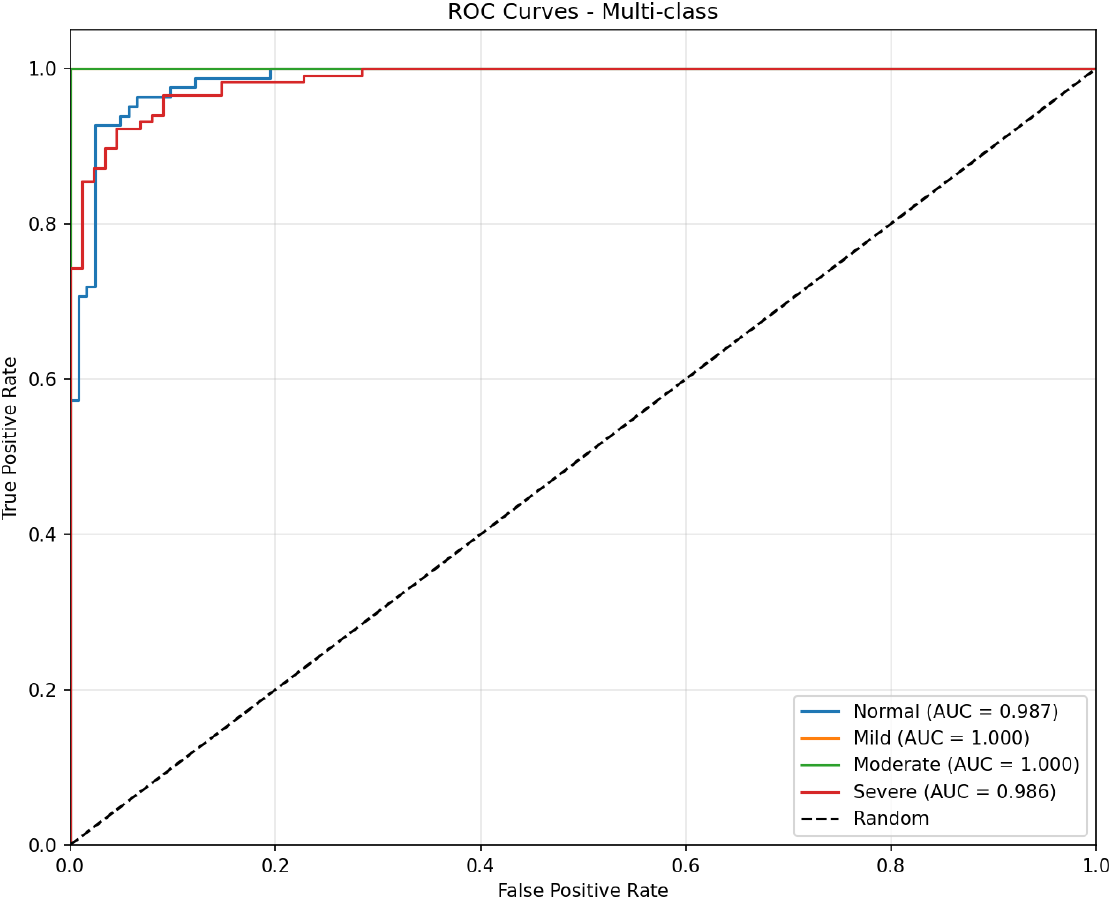
Per-class ROC curves for the ensemble on the test set.

## 5 Discussion

The proposed framework advances sFGR detection by combining GA-independent labeling, robust abdominal circumference estimation, and a severity-aware deep learning classifier. From a methodological standpoint, the results indicate that carefully designed preprocessing and feature extraction can compensate for the absence of explicit gestational age information and still achieve performance that is comparable to or better than recent state-of-the-art approaches that rely on richer input modalities [Dong et al., 2025, Mikołaj et al., 2025].

The test set performance, with 93.7% accuracy, weighted F1-score of 0.76, and AUC values above 0.98 for all classes, suggests that the model captures clinically relevant patterns in fetal abdominal morphology. Comparison with Dong et al. [2025], who reported an F1-score of 0.765 and AUC of 0.80 for early FGR using placental images, indicates that the proposed approach at least matches, and in some aspects exceeds, prior work while using standard 2D abdominal ultrasound. Similarly, the SGA detection sensitivity reported by likołaj et al. [2025] of 70% with their deep learning model trained on a substantially larger dataset suggests that complementary approaches to fetal growth assessment are converging on clinically useful performance levels, although direct numerical comparison is limited by differences in study design and endpoints.

From a clinical perspective, the model is best interpreted as a first-line screening and triage tool rather than a replacement for guideline-recommended FGR work-up. In a typical workflow, low-risk predictions would reassure providers that immediate escalation may not be needed, whereas high-risk predictions would trigger referral for a full biometry and Doppler assessment that incorporates head circumference, femur length, uterine and umbilical artery Doppler indices, and maternal risk factors. Restricting the input to a single abdominal plane mirrors what can realistically be acquired by mid-level providers on basic machines, and is therefore a deliberate trade-off in favour of deployability in low-resource clinics.

A key modelling decision was the use of percentile-based proxy labels derived from abdominal circumference at term, rather than true EFW or neonatal outcomes. This choice reflects the constraints of the public dataset and is aligned with widely used FGR definitions that operationalize severity in terms of GA-adjusted EFW or AC percentiles [Chew and Verma, 2023, Tongsong et al., 1999, Stirnemann et al., 2020]. Anchoring all cases to a common 37-week reference frame allows us to obtain a gestational-agenormali1ed size ranking that can be learned from images alone. The main risk is that these proxy labels may misclassify constitutionally small but healthy fetuses or fail to capture growth-restricted fetuses with relatively preserved AC. In future work, we plan to perform ablation studies in cohorts with reliable GA, EFW, and outcome data to estimate the magnitude and direction of this bias and to refine the proxy accordingly.

For screening applications in LMICs, high NPV is particularly important, as it supports safe exclusion of low-risk pregnancies from further resource-intensive evaluation. In the present framework, strong performance on the normal class and the pattern of errors concentrated between neighboring severity levels both support this use case. Severe sFGR cases are rarely misclassified as normal, which reduces the risk of missed high-risk pregnancies while still flagging a manageable number of cases for Doppler and specialist review.

Beyond pure diagnostic performance, the framework also has practical advantages. It operates on single 2D abdominal images and can be integrated into existing ultrasound workflows without additional sensors or substantial modifications of acquisition protocols. The DenseNet-121 backbone can be quantized and deployed on mid-range GPUs or even on modern CPUs with optimized inference engines, which is compatible with hardware available in many tertiary centers and some secondary-level facilities in LMICs.

Despite these strengths, several limitations must be acknowledged, which also shape how the results should be interpreted. First, the dataset originates from a single tertiary center in Brazil. As highlighted by recent work on the generalizability of fetal ultrasound deep learning models across African sites [Sendra-Balcells et al., 2023], performance can drop substantially when models are transported between scanners, acquisition protocols, and populations. Our results should therefore be viewed as a proof-of-concept demonstration on this specific dataset rather than as evidence of ready-to-deploy global generalizability. Second, the number of samples per intermediate severity category is modest, which likely contributes to the lower cross-validation AUC values observed for mild and moderate classes. Third, the severity labels are derived from percentile-based proxies rather than true birth-weight or perinatal outcomes, and depend on the quality of the abdominal masks used to compute AC. The observed discrepancy between the tails of the perimeter- and ellipse-based AC distributions underscores the importance of explicit quality control and motivates future feature-importance and ablation analyses to quantify how sensitive model performance is to each measurement choice.

The current work also focuses exclusively on static 2D abdominal images. Dynamic information from cine loops, Doppler studies of the umbilical and middle cerebral arteries, or complementary biometric measurements such as head circumference and femur length could further refine severity stratification. Temporal models uncertainty quantification, and self-supervised pretraining on large unlabeled ultrasound datasets are promising directions to improve robustness and interpretability.

Future work should therefore prioritize multi-site validation with datasets from diferent continents and healthcare systems, including prospective evaluation in LMIC clinics. Integrating the model into a human-in-the-loop workflow, where high-confidence predictions inform but do not replace expert judgment, may be a pragmatic path towards clinical adoption. Edge deployment on portable ultrasound devices and telemedicine platforms could extend the reach of specialized obstetric expertise to rural and underserved populations. In parallel, systematic ablation of alternative AC measurement strategies and comparison of proxy labels with outcome-based definitions will help clarify which design choices are most critical for clinical performance.

## 6 Conclusion

This study presented a GA-independent deep learning framework for the screening and severity classification of fetal growth restriction using 2D fetal abdominal ultrasound images. By combining a dual-method abdominal circumference estimation pipeline with a DenseNet-121-based classifier and patient-wise ensemble evaluation, the framework achieved high accuracy, strong class-wise performance, and excellent discriminative ability on a held-out clinical test set.

The results indicate that standard 2D ultrasound, when coupled with modern deep learning techniques, can support reliable, severity-aware sFGR screening in settings where precise gestational dating and advanced imaging resources are limited. While further validation is needed in multi-center and prospective cohorts, the proposed approach represents a step towards more equitable prenatal care, with the potential to act as a triage layer that reduces preventable morbidity and mortality associated with fetal growth restriction in low-resource environments.

## Data Availability

The Fetal Abdominal Structures Segmentation Dataset used in this study is publicly available [Da Correggio et al., 2024]. Code and trained models will be made available upon reasonable request to the corresponding author.

## Ethics Statement

This study used a publicly available, de-identified dataset. Ethical approval for the original data collection was obtained from the UFSC ethics committee as described by Da Correggio et al. [2024].

## Author Contributions

A.E. conceived the study, designed the framework, did literature review, designed the methodology, implemented the models, performed the experiments, and wrote the manuscript. R.A.A. contributed to data analysis and manuscript revision. All authors reviewed and approved the final manuscript.

## Conflict of Interest

The authors declare no competing interests.

## Funding

This research received no external funding.

## Notes

### Competing Interest Statement

The authors have declared no competing interest.

### Clinical Trial

4.971.754

### Author Declarations

This study used a publicly available, de-identied dataset. Ethical approval for the original data collection was obtained from the UFSC ethics committee as described by Da Correggio et al. [2024].

